# Post-acute COVID-19 syndrome negatively impacts health and wellbeing despite less severe acute infection

**DOI:** 10.1101/2020.11.04.20226126

**Authors:** Laura Tabacof, Jenna Tosto-Mancuso, Jamie Wood, Mar Cortes, Amy Kontorovich, Dayna McCarthy, Dahlia Rizk, Nicki Mohammadi, Erica Breyman, Leila Nasr, Christopher Kellner, David Putrino

## Abstract

**Introduction:** One of the noted features of COVID-19 is the spectrum of expressivity in symptoms among those with the disease, ranging from no or mild symptoms that may last a small number of days, to severe and/or longer lasting symptoms. It is emerging that many patients have long lasting symptoms, several months after initial infection with COVID-19. The aim of this research was to characterize post-acute COVID-19 syndrome (PACS).

**Methods:** This was a retrospective cross-sectional observational study. Participants were patients recovering from COVID-19 infection, enrolled in Mount Sinai Hospital’s COVID-19 Precision Recovery Program (PRP). Inclusion criteria were confirmed or probable (based on World Health Organization criteria) initial diagnosis of COVID-19; post-acute COVID-19 syndrome (defined as experiencing symptoms > 6 weeks since acute symptom onset) and being currently enrolled in the PRP during the months of July and August 2020. Study survey data were collected using REDCap. Demographic data, COVID-19 clinical data and patient-reported outcomes for breathlessness (Medical Research Council Breathlessness Scale), fatigue and quality of life (EuroQoL 5D-5L) were collected.

**Results:** 84 individuals with PACS were included. Symptoms persisted at mean (range) 151 (54 to 255) days. The most prevalent persistent symptoms were fatigue (92%), loss of concentration/memory (74%), weakness (68%), headache (65%) and dizziness (64%). Most participants reported increased levels of disability associated with breathlessness, increased fatigue and reduced quality of life.

**Conclusions:** Persistent symptoms following COVID-19 infection are prevalent, debilitating and appear to affect individuals regardless of acute infection severity or prior health status. More detailed research is required in order to identify specific symptom clusters associated with PACS, and to devise effective interventional strategies.

## Introduction

One of the noted features of COVID-19 is the spectrum of expressivity in symptoms among those with the disease, ranging from no or mild symptoms that may last a small number of days, to severe and/or longer lasting symptoms [1, 2]. Severe acute COVID-19 symptoms can coincide with respiratory failure and potentially death, or ongoing medical problems [3, 4]. The symptomatology is complex [5, 6], and further work is required to describe the presence and impact of longer lasting, or “post-acute” symptoms.

Symptom variability, especially in those with post-acute COVID-19 syndrome (PACS), presents challenges to healthcare teams and patients. There are difficulties associated with devising and delivering treatment programs amidst such disease novelty and diversity, which can be a source of frustration for both clinicians and patients with COVID-19 alike. Moreover, the majority of current healthcare efforts center on prevention and acute management [7, 8]. As a result, there remains a limited understanding of, and resource allocation to, patients who continue to display debilitating symptoms weeks and months following initial diagnosis. To date, the presence of persistent symptoms months after initial COVID-19 diagnosis has only been described in brief, and only in those whose acute illness was severe [9].

This study describes the detailed symptom data obtained from patients with PACS, and the impact of these symptoms on self-reported physical function, quality of life and wellbeing. The aim of this study was to characterise the persistent symptoms experienced by this cohort, and to better inform healthcare provision for PACS.

## Materials and methods

This was a cross-sectional observational study. Approval for the use of Precision Recovery Program (PRP) data was provided by the Mount Sinai Program for Protection of Human Subjects (IRB 20-03315).

### Participants

Participants were patients with past COVID-19 infection, referred to Mount Sinai Hospital’s COVID-19 Precision Recovery Program (PRP) by a medical doctor, either during the acute or post-acute phase of their disease. Inclusion criteria were confirmed or probable (i.e. confirmed by a medical doctor in accordance with World Health Organization recommendations [WHO] [10]) initial diagnosis of COVID-19; PACS (defined as experiencing symptoms > 6 weeks since initial symptom onset); and provision of consent for clinical data to be used for research purposes.

### Data collection and Outcomes

Study data were collected and managed using REDCap (Research Electronic Data Capture) electronic data capture tools hosted at Mount Sinai Health System. REDCap is a secure, web-based application designed to support data capture for research studies [11]. Survey questions used within REDCap were developed or selected by the Mount Sinai PRP team, including components developed in conjunction with the WHO COVID-19 Technical Working Group. Participants were provided with a link to the survey via email.

Baseline demographic data included gender, age, body mass index (BMI) and comorbidities. COVID-19 clinical data included date of acute COVID-19 symptom onset, polymerase chain reaction (PCR) and antibody test results, need for hospitalization and oxygen supplementation, and acute and persistent symptoms. Patient-reported outcomes included screening tools for breathlessness (Medical Research Council [MRC] Breathlessness Scale) [12], fatigue (visual analogue scale) and quality of life (QoL) (EuroQol EQ-5D-5L) [13]. Participants were also asked to rate whether their QoL was the same, better or worse following contracting COVID-19.

### Statistical analyses

Statistical analyses were undertaken with Stata (StataCorp, Stata Statistical Software Release: V.14). Data were analysed using descriptive statistics.

## Results

Data is reported on the first 84 participants who met the inclusion criteria and responded to the survey (Table 1). Eighty-two (98%) participants reported they underwent clinical testing for COVID-19 via PCR and/or antibody tests, with 33 (39%) positive for the disease on at least one test (Table1). The most prevalent symptoms experienced during acute COVID-19 infection were dyspnoea (82%), fever/chills (79%), tinnitus (75%), loss of concentration/memory (73%) and weakness (70%) (Figure 1).

**Table 1.**
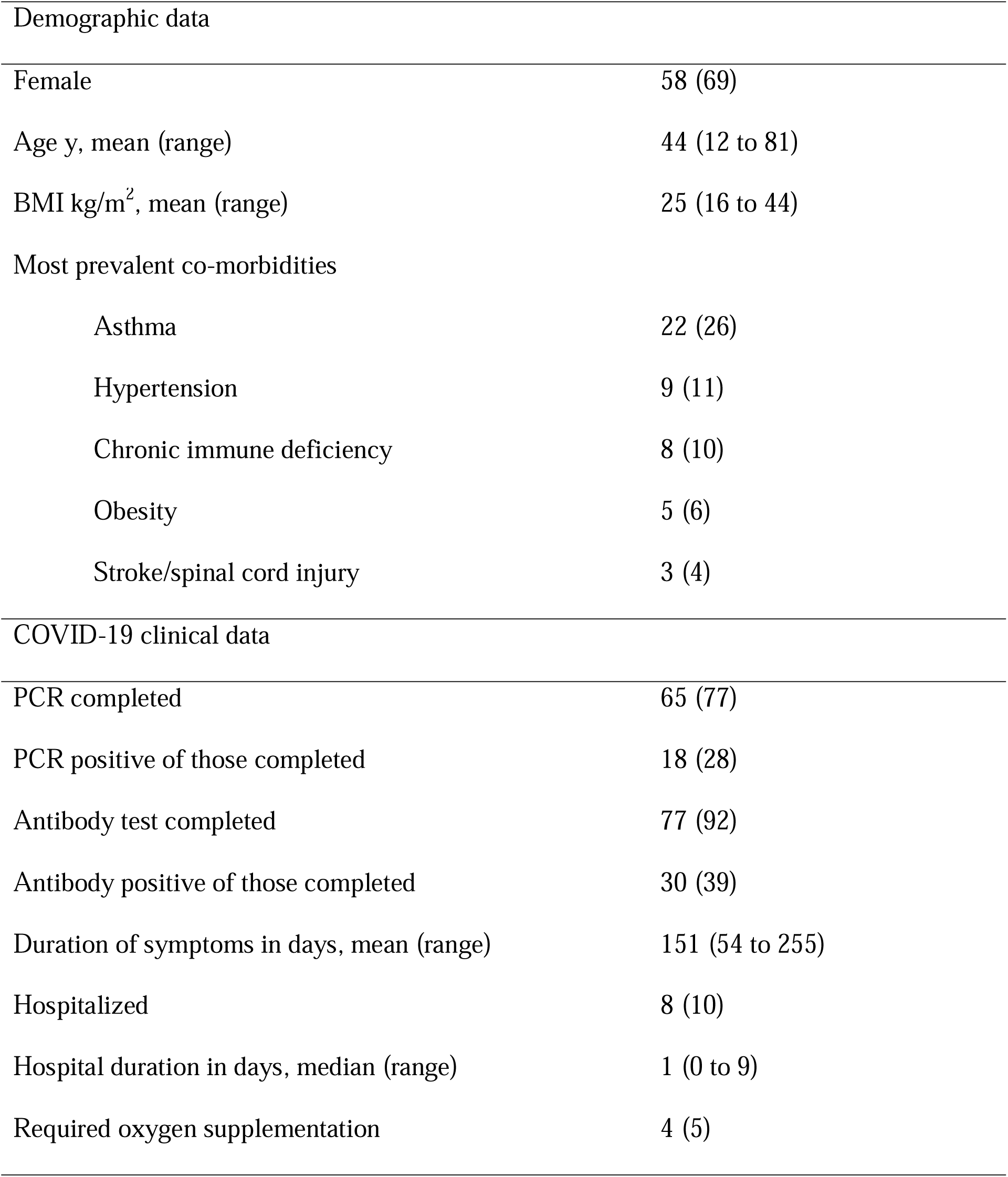
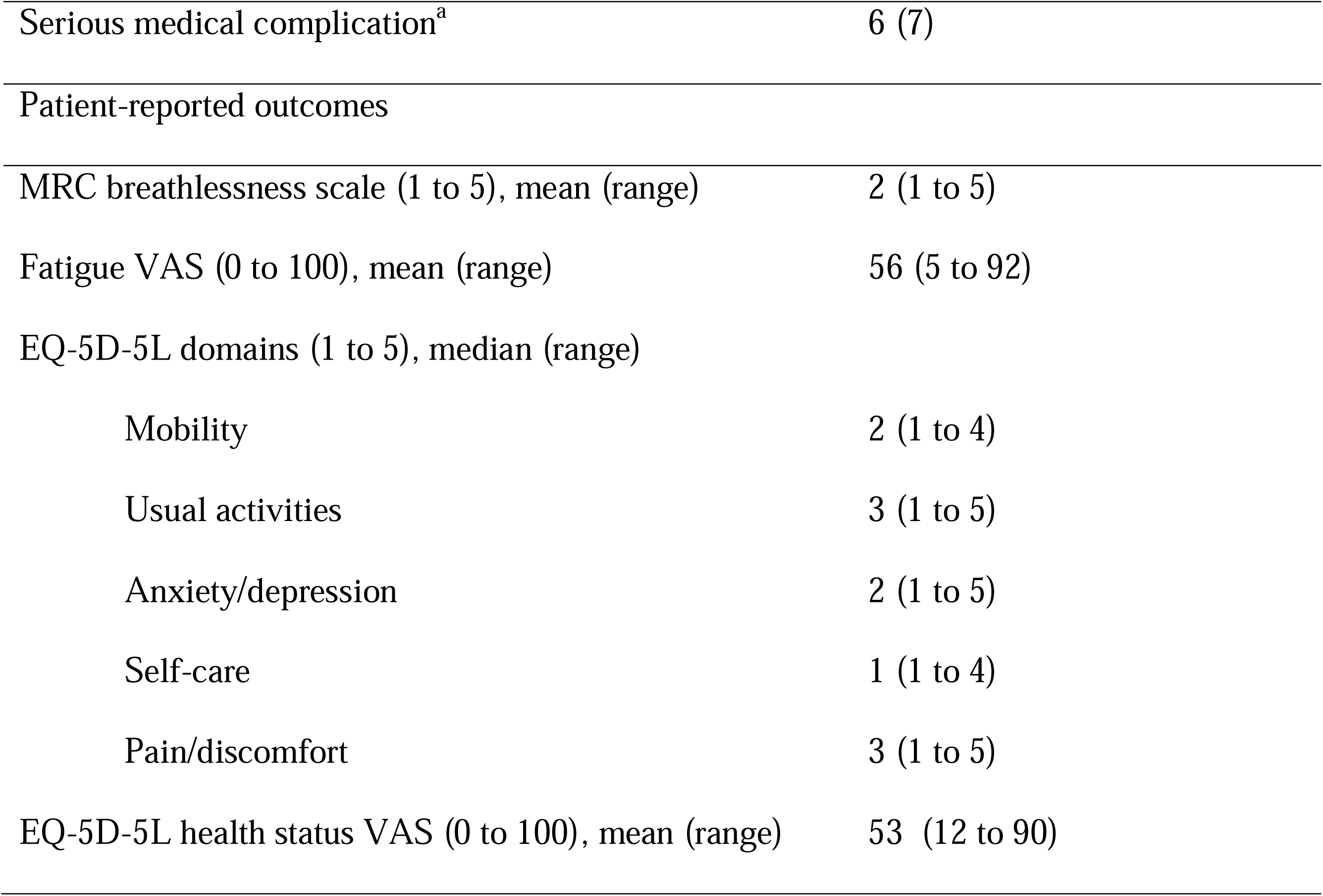
Participant (n=84) baseline demographic, COVID-19 clinical data and patient-reported outcomes. Data are presented as n (%) unless otherwise indicated. aSerious medical complications reported included stroke, kidney and liver dysfunction. MRC = Medical Research Council; VAS = visual analogue scale.

**Figure 1.**
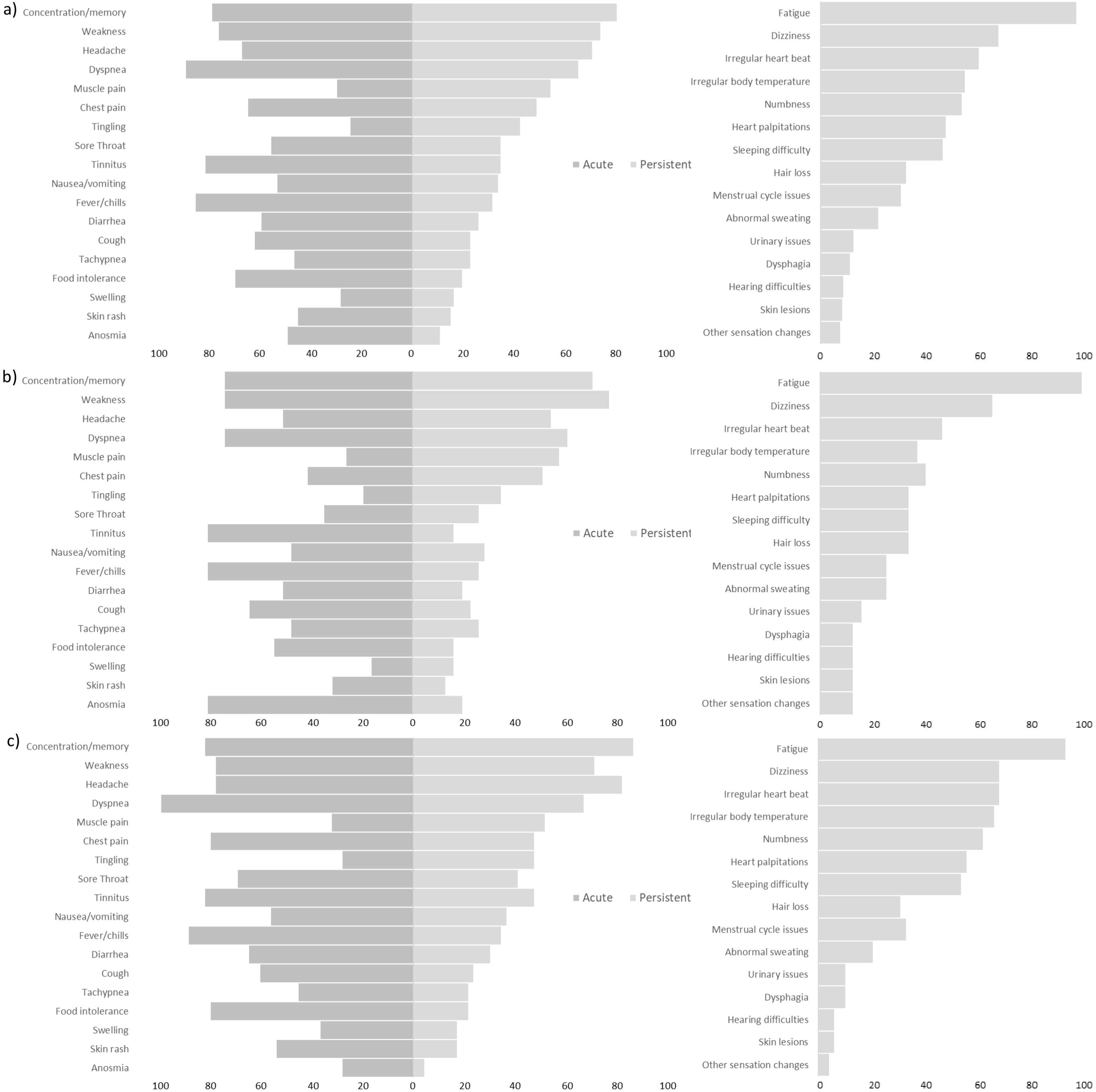
Symptoms reported by participants in the acute and persistent phases, including additional persistent symptoms: a) all participants (n=84); b) participants with confirmed COVID-19 via PCR or antibody test (n=34); c) presumptive positive COVID-19 (n=50).

Participants reported persistent symptoms at mean (range) 151 (54 to 255) days. Thirty-five symptoms were reported among patients in the persistent phase, (all 19 acute symptoms plus 16 additional symptoms). The most prevalent persistent symptoms reported were fatigue (92%), loss of concentration/memory (74%), weakness (68%), headache (65%) and dizziness (64%) (Figure 1). Patterns of persistent symptoms were similar in participants when divided into two groups, being confirmed (via PCR and/or antibody tests) and probable COVID-19 infection (Figure 1).

The majority of participants (88%) reported increased levels of disability associated with breathlessness on the MRC breathlessness scale (grades 2 and above). The mean (range) level of fatigue reported was 56 (5 to 92) out of 100, with two thirds (65%) reporting fatigue levels of ≥ 50/100. The domains of the EQ5D5L with the highest scores were ‘usual activities’ and ‘pain/discomfort’ (Table 1). Most participants (82%) reported that their QoL was worse now compared to before contracting COVID-19.

## Discussion

Post-acute COVID-19 syndrome is a collection of symptoms lingering long after acute infection. Participants in this study reported a significant burden of symptomatology impacting their self-reported physical function, quality of life and wellbeing.

The data presented in this study highlight the prevalence of persistent and debilitating symptoms in a cohort of patients with previous confirmed or presumed COVID-19 infection that was predominantly managed without the need for hospitalisation. This work builds upon the work of Carfi et al (9) by demonstrating that serious sequelae are not limited to individuals who experienced severe acute COVID-19 infection. Increased levels of fatigue, breathlessness and weakness were common and likely affected the QoL scores observed on the EQ5D5L, in particular relating to the ‘usual activities’ and ‘pain/discomfort’ domains.

The prevalence of these symptoms in previously healthy, relatively young adults, identifies long-term sequelae that are associated with even less severe cases of COVID-19 infection. Although the proportion of positive PCR and/or antibody tests was 39%, this reflects concerns raised regarding the accuracy and potentially high rate of false-negatives seen in testing [14, 15, 16], the limited access to testing earlier in the pandemic, and participants possibly delaying seeking PCR tests during acute infection. The diagnosis of probable COVID-19 infection is supported by the similarity in persistent symptom presentation shared with those who received positive COVID-19 tests.

Long-lasting symptoms not yet defined by any current diagnostic paradigm can be extremely daunting for patients and give rise to a sense of isolation. As such, clinicians are urged to acknowledge the presence of the persistent symptoms that can impact the health and wellbeing of patients with PACS.

Persistent symptoms following COVID-19 infection are prevalent, debilitating and appear to affect patients regardless of acute infection severity or prior health status. It is more important than ever to promote suppression and prevention strategies as more is understood about the potential for COVID-19 infection to give way to long-term health effects in healthy individuals. More detailed research is required in order to identify specific symptom clusters and predictors associated with PACS, and to devise effective interventional strategies.

## Data Availability

The authors confirm that all data underlying the findings described in their manuscript are available upon request

## Acknowledgements

The authors would like to acknowledge the participants in the study, the frontline healthcare workers at Mount Sinai Health System, and the wider research team at the Abilities Research Center and the Center for Post-COVID Care at Mount Sinai.

## Supplementary material

